# Sociodemographic inequality in COVID-19 vaccination coverage amongst elderly adults in England: a national linked data study

**DOI:** 10.1101/2021.05.13.21257146

**Authors:** Vahé Nafilyan, Ted Dolby, Cameron Razieh, Charlotte Gaughan, Jasper Morgan, Daniel Ayoubkhani, A. Sarah Walker, Kamlesh Khunti, Myer Glickman, Thomas Yates

## Abstract

**Objective:** To examine inequalities in COVID-19 vaccination rates amongst elderly adults in England

**Design:** Cohort study

**Setting:** People living in private households and communal establishments in England

**Participants:** 6,829,643 adults aged ≥ 70 years (mean 78.7 years, 55.2% female) who were alive on 15 March 2021.

**Main outcome measures:** Having received the first dose of a vaccine against COVID-19 by 15 March 2021. We calculated vaccination rates and estimated unadjusted and adjusted odds ratios using logistic regression models.

**Results:** By 15 March 2021, 93.2% of people living in England aged 70 years and over had received at least one dose of a COVID-19 vaccine. While vaccination rates differed across all factors considered apart from sex, the greatest disparities were seen between ethnic and religious groups. The lowest rates were in people of Black African and Black Caribbean ethnic backgrounds, where only 67.2% and 73.9% had received a vaccine, with adjusted odds of not being vaccinated at 5.01 (95% CI 4.86 - 5.16) and 4.85 (4.75 - 4.96) times greater than the White British group. The proportion of individuals self-identifying as Muslim and Buddhist who had received a vaccine was 79.1% and 84.1%, respectively. Older age, greater area deprivation, less advantaged socio-economic position (proxied by living in a rented home), being disabled and living either alone or in a multi-generational household were also associated with higher odds of not having received the vaccine.

**Conclusion:** People disproportionately affected seem most hesitant to COVID-19 vaccinations. Policy Interventions to improve these disparities are urgently needed.

**Summary Box:** *What is already known on this subject?:* The UK began an ambitious vaccination programme to combat the COVID-19 pandemic on 8th December 2020. Existing evidence suggests that COVID-19 vaccination rates differ by level of area deprivation, ethnicity and certain underlying health conditions, such as learning disability and mental health problems.

*What does this study add?:* Our study shows that first dose vaccination rates in adults aged 70 or over differed markedly by ethnic group and self-reported religious affiliation, even after adjusting for geography, socio-demographic factors and underlying health conditions. Our study also highlights differences in vaccination rates by deprivation, household composition, and disability status, factors disproportionately associated with SARS-CoV-2 infection. Public health policy and community engagement aimed at promoting vaccination uptake is these groups are urgently needed.

*Strengths and limitations of this study:*

- Using nationwide linked population-level data from clinical records and the 2011 Census, we examined a wide range of socio-demographic characteristics not available n electronic health records
- Most demographic and socio-economic characteristics are derived from the 2011 Census and therefore are 10 years old. However, we focus primarily on characteristics that are unlikely to change over time, such as ethnicity or religion, or likely to be stable for our population
- Because the data are based on the 2011 Census, it excluded people living in England in 2011 but not taking part in the 2011 Census; respondents who could not be linked to the 2011-2013 NHS patients register; recent migrants. Consequently, we excluded 5.4% of vaccinated people who could not be linked

## Introduction

The UK began an ambitious vaccination programme to combat the COVID-19 pandemic on 8th December 2020; by 24th April 2021, 64% of the UK adult population have received their first of the dose [1].

Previous research demonstrates that vaccination rates tend to be lower amongst certain ethnic groups, and in areas of higher deprivation [2, 3, 4]. Existing evidence suggests that COVID-19 vaccination rates differ by level of area deprivation, certain underlying health conditions and ethnicity [5]. Far less is known about how COVID-19 vaccination uptake varies by socio-demographic factors, such as religious affiliation, individual socio-economic status, living in multigenerational household or disability status, factors disproportionately associated with SARS-CoV-2 infection. Understanding which socio-demographic, economic, and cultural factors are associated with low vaccination rates has major implications for designing policies that help maximise the vaccination campaign coverage.

This study investigates inequality in vaccination rates amongst adults aged ≥ 70 years in England, using population-level administrative records linked to the 2011 Census. This enables examination of a wide range of socio-demographic characteristics, currently lacking in previously published studies, in particular ethnicity, religion, different measures of socio-economic position, and those who report being disabled.

## Methods

We linked vaccination data from the National Immunisation Management System (NIMS) to the Office for National Statistics (ONS) Public Health Data Asset (PHDA) based on NHS number. The ONS PHDA is a linked dataset combining the 2011 Census, mortality records, the General Practice Extraction Service (GPES) data for pandemic planning and research and the Hospital Episode Statistics (HES). To obtain NHS numbers for the 2011 Census, we linked the 2011 Census to the 2011-2013 NHS Patient Registers using deterministic and probabilistic matching, with an overall linkage rate of 94.6%. All subsequent linkages were performed based on NHS numbers.

The study population consisted of people aged ≥ 70 years, alive on 15th March 2020, who were resident in England, registered with a general practitioner, and enumerated at the 2011 Census. Of 6,605,315 adults aged ≥ 70 years who received a first dose of a COVID-19 vaccine in NIMS, 6,242,384 (94.5%) were linked to the ONS PHDA.

The main outcome was having received at least a first dose of a COVID-19 vaccine by 15th March 2021, as recorded in the NIMS data available on 31st March 2021. Phase 1 of the vaccination policy for England aimed to offer a first vaccination appointment to all those ≥ 70 years by 15th February, and we allowed a further month to ensure full coverage.

This dataset combines comprehensive socio-demographic information from the 2011 Census with a detailed medical history from clinical records. All individual-level socio-demographic characteristics (ethnic group, religious affiliation, disability status, educational attainment) came from the 2011 Census. We used a 10-category ethnic group classification (white British, Bangladeshi, black African, black Caribbean, Chinese, Indian, Mixed, Other, Pakistani, white other). Self-reported religious group, place of residence (region within England, private or care home) and area-based deprivation (Index of Multiple Deprivation [6]) were derived based on the 2019 Patient Register. Comorbidities were defined as in the QCOVID risk prediction model [7]. All variables included in this analysis are listed in Supplementary Table S1.

First, we estimated the first dose vaccination rates by a range of demographic and socio-economic characteristics. Second, to understand the drivers of the observed differences in vaccination rates, we used logistic regression to estimate the odds of not having received a first dose of a COVID-19 vaccine. For each exposure, we compared odds ratios from an unadjusted model, a model only adjusted for sex and age, and a model further adjusted for all geographic and socio-demographic characteristics, disability status and pre-existing conditions.

Patient and Public Involvement

No patient involved

## Results

Our study population included 6,724,179 adults aged ≥ 70 years who lived in England. 55.1% were women and the mean age was 78.8 (SD: 6.5) years; 91.6% identified themselves as white British, 78.5% as Christian. 82.5% owned their home (Table 1). By 15 March 2021, 93.2% of people living in England aged 70 years and over had received at least one dose of a COVID-19 vaccine.

**Table 1:**
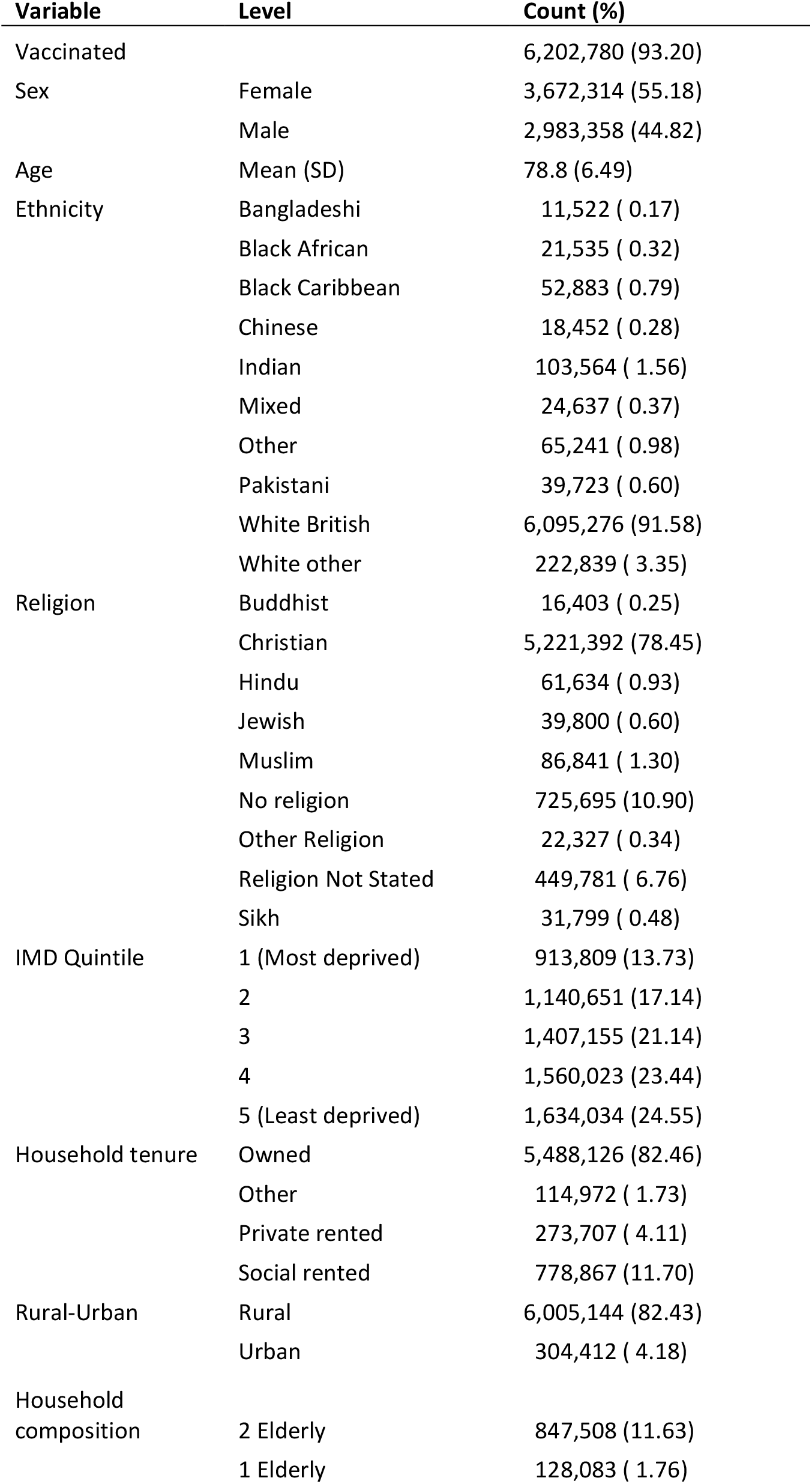
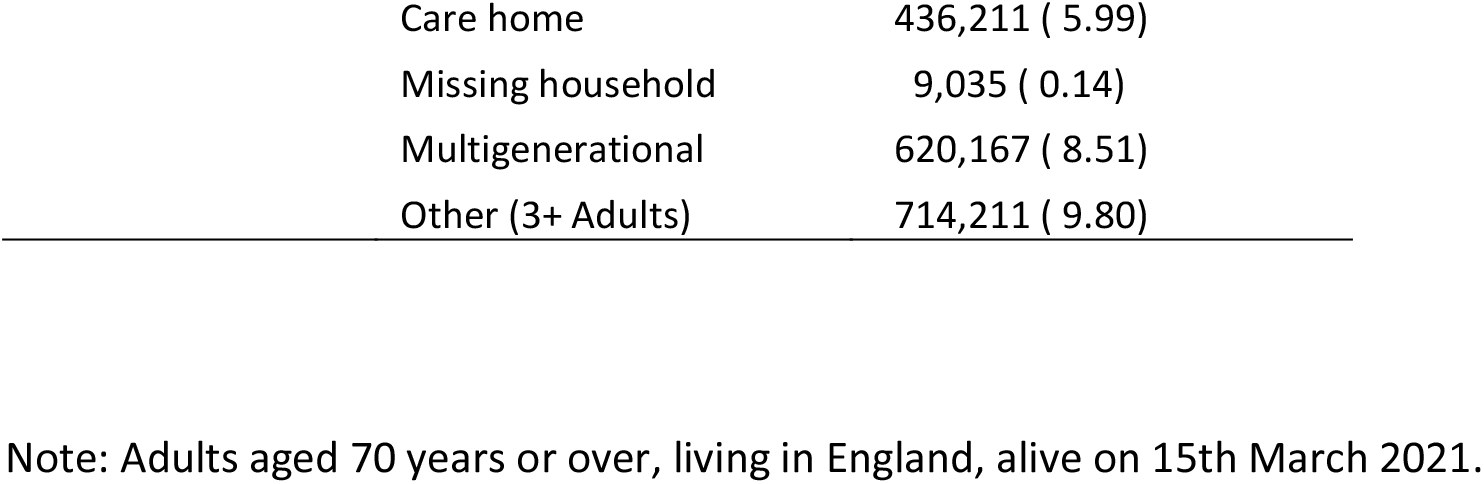
Characteristics of the study population

| Variable | Level | Count (%) |
| --- | --- | --- |
| Vaccinated |  | 6,202,780 (93.20) |
| Sex | Female | 3,672,314 (55.18) |
|  | Male | 2,983,358 (44.82) |
| Age | Mean (SD) | 78.8 (6.49) |
| Ethnicity | Bangladeshi | 11,522 ( 0.17) |
|  | Black African | 21,535 ( 0.32) |
|  | Black Caribbean | 52,883 ( 0.79) |
|  | Chinese | 18,452 ( 0.28) |
|  | Indian | 103,564 ( 1.56) |
|  | Mixed | 24,637 ( 0.37) |
|  | Other | 65,241 ( 0.98) |
|  | Pakistani | 39,723 ( 0.60) |
|  | White British | 6,095,276 (91.58) |
|  | White other | 222,839 ( 3.35) |
| Religion | Buddhist | 16,403 ( 0.25) |
|  | Christian | 5,221,392 (78.45) |
|  | Hindu | 61,634 ( 0.93) |
|  | Jewish | 39,800 ( 0.60) |
|  | Muslim | 86,841 ( 1.30) |
|  | No religion | 725,695 (10.90) |
|  | Other Religion | 22,327 ( 0.34) |
|  | Religion Not Stated | 449,781 ( 6.76) |
|  | Sikh | 31,799 ( 0.48) |
| IMD Quintile | 1 (Most deprived) | 913,809 (13.73) |
|  | 2 | 1,140,651 (17.14) |
|  | 3 | 1,407,155 (21.14) |
|  | 4 | 1,560,023 (23.44) |
|  | 5 (Least deprived) | 1,634,034 (24.55) |
| Household tenure | Owned | 5,488,126 (82.46) |
|  | Other | 114,972 ( 1.73) |
|  | Private rented | 273,707 ( 4.11) |
|  | Social rented | 778,867 (11.70) |
| Rural-Urban | Rural | 6,005,144 (82.43) |
|  | Urban | 304,412 ( 4.18) |
| Household composition | 2 Elderly | 847,508 (11.63) |
|  | 1 Elderly | 128,083 ( 1.76) |

|  |  |
| --- | --- |
| Care home | 436,211 ( 5.99) |
| Missing household | 9,035 ( 0.14) |
| Multigenerational | 620,167 ( 8.51) |
| Other (3+ Adults) | 714,211 ( 9.80) |

Note: Adults aged 70 years or over, living in England, alive on 15th March 2021.

Table 2 shows vaccination rates by demographic and socio-economic factors, as well as odds ratios (OR) from different models. Vaccination rates differed across all factors considered, apart from sex. The lowest rates were in people of Black African and Black Caribbean ethnic backgrounds where only 67.2% and 73.9% had received a vaccine. Adjusting for differences in geography, socio-demographic factors and underlying health conditions did not fully explain the lower probability of having received the vaccine among ethnic minority groups. Compared with people of white British ethnicity, the fully adjusted OR for black African individuals was 5.01 [95% CI 4.86 - 5.16], whilst the unadjusted OR was 7.62 [7.40 - 7.84], suggesting that geography, socio-demographic factors and pre-pandemic health only explains about 40% of the elevated odds of not being vaccinated.

**Table 2.**
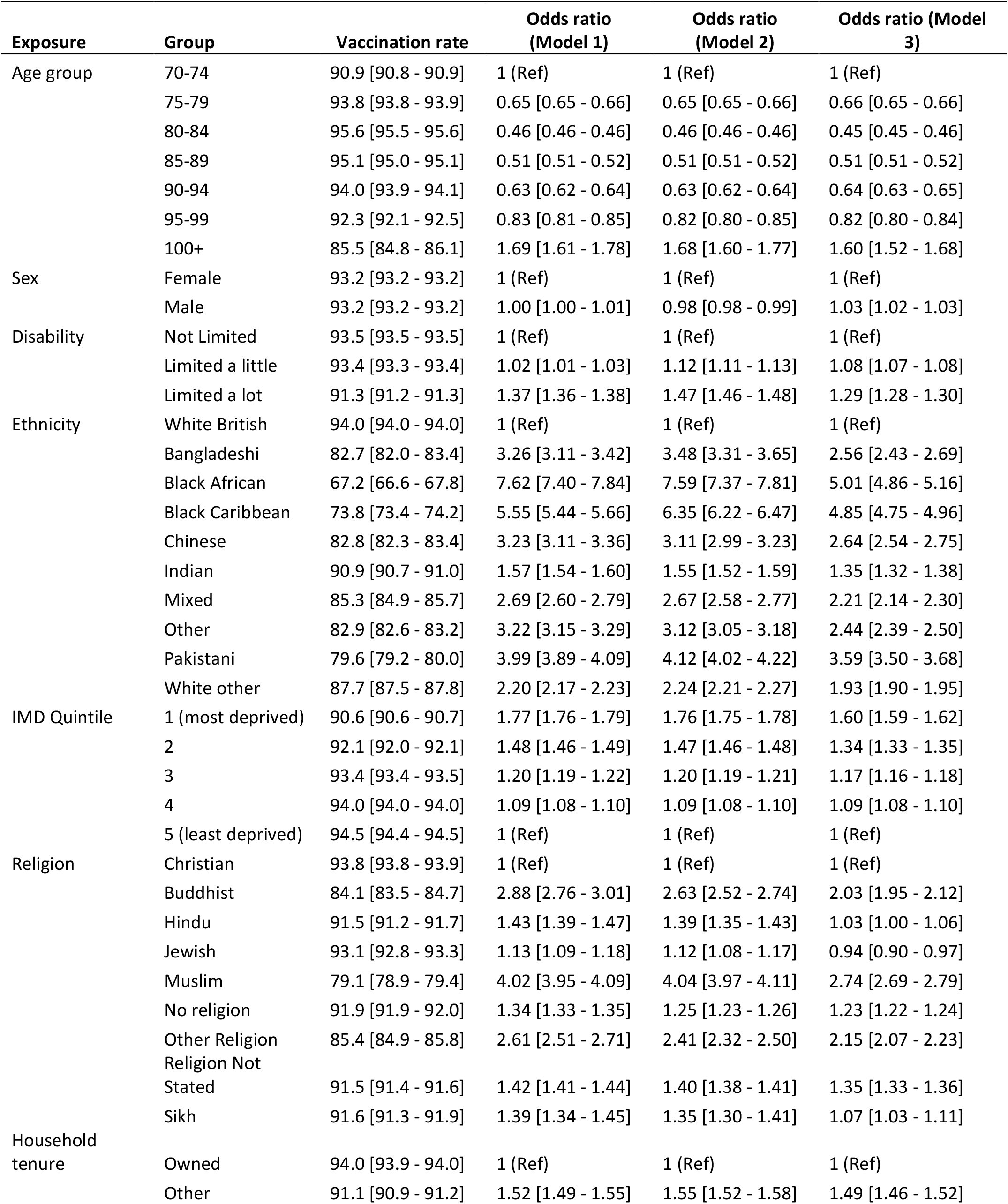
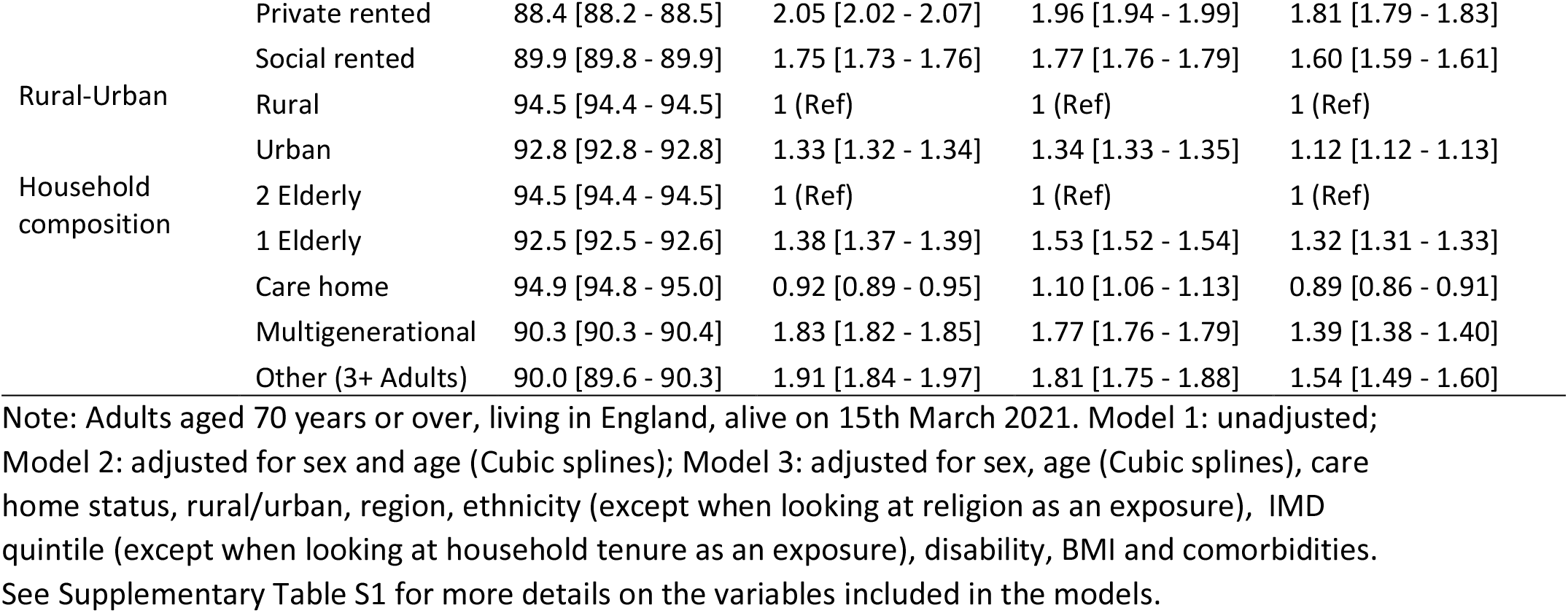

Vaccination rates also varied markedly across religious groups. Whilst 93.8% of Christians had been vaccinated, only 79.1% of Muslims and 84.1% of Buddhists had been vaccinated. Stark differences remained after adjustment for other factors, with an adjusted OR of not being vaccinated of 2.74 [2.69 - 2.79] for Muslims and 2.03 [1.95 - 2.12] for Buddhist, compared to Christians.

Greater area deprivation, less advantaged socio-economic position (proxied by living in a rented home), being disabled and living either alone or in a multi-generational household were also associated with low vaccination rates, even when adjusting for other factors (Table 2). These differences were less pronounced than the differences between ethnic groups or religious affiliations.

## Discussion

### Main findings

Our analysis using whole population level linked data in England suggests that first dose vaccination rates in adults aged ≥ 70 differed markedly by ethnic group and self-reported religious affiliation. The percentage of people vaccinated was lower among all minority ethnic groups compared to the White British population, with the lowest vaccination rates observed among Black African, Black Caribbean, Bangladeshi and Pakistani individuals. In addition, lower vaccination rates were reported among individuals who identified as Muslim and Buddhist. Whilst some differences were found by deprivation, household factors, disability status and other socio-demographic factors, these were less pronounced compared with ethnicity or religious affiliation.

### Comparison with other studies

Few studies have investigated how COVID-19 vaccination coverage varies by a wide range of socio-demographic characteristics. Our results on ethnicity and area deprivation are consistent with one previous study based on clinical records for 40% of patients in England [3]. In addition, our results confirm studies showing that influenza, shingles and pneumococcal vaccination are patterned by similar factors, including ethnicity, deprivation and household size [8]. Pre-pandemic, religion and culture have been postulated to be important factors in determining vaccination uptake [9]; our results extend this by showing that self-reported religious affiliation is an important factor in COVID-19 vaccine uptake. Differences in vaccination rate and potential vaccination hesitancy between religious groups may not be based upon religious beliefs, but rather reflect safety and other concerns [10], or, given high infection rates in some of these groups [11], beliefs that vaccination is not needed after natural infection. We also find that vaccination rates vary by individual characteristics not reported in previous studies, such as household tenure (a proxy for socio-economic status), household composition and disability status.

### Strengths and limitation

The primary study strength is using nationwide linked population-level data from clinical records and the 2011 Census. Unlike studies based solely on electronic health records, we examined a wide range of socio-demographic characteristics. Unlike surveys, we can precisely estimate vaccination rates and odds ratios for small groups. The main limitation is that most demographic and socio-economic characteristics are derived from the 2011 Census and therefore are 10 years old. However, we focus primarily on characteristics that are unlikely to change over time, such as ethnicity or religion, or likely to be stable for our population (adults aged ≥ 70), such as household tenure. However, for the characteristics likely to change over time, such as disability status, the time difference may introduce some bias into the estimates, although this would be expected to dilute differences, since we are most likely missing some long-term health conditions. Care home residency and area deprivation were derived from the 2019 Patient register and are therefore not subject to the same biases. Another limitation is that because the Public Health Data Asset was based on the 2011 Census, it excluded people living in England in 2011 but not taking part in the 2011 Census; respondents who could not be linked to the 2011-2013 NHS patients register; recent migrants. Consequently, we excluded 5.4% of vaccinated people who could not be linked to the ONS PHDA.

## Conclusion

As ethnicity and religion may be the most important determinants of COVID-19 vaccination rates in England, research is now urgently needed to understand why these disparities exist in these groups and how they can best be addressed through public health policy and community engagement. This is especially important because the groups with low vaccination coverage were also at elevated risk of COVID-19 mortality in the first two waves of the pandemic [12, 11, 13, 14].

## Data Availability

The ONS Public Health Linked Data Asset will be made available on the ONS Secure Research Service for Accredited researchers. Researchers can apply for accreditation through the Research Accreditation Service.

## Acknowledgements

We are grateful to Charlotte Bermingham for useful discussions about this paper.

## Ethics approval

Ethical approval was obtained from the National Statistician’s Data Ethics Advisory Committee (NSDEC(20)12)

## Appendix

**Supplementary Table 1:** Variables used in the analyses

| Variable | Coding | Source |
| --- | --- | --- |
| Vaccinated | Received a first dose of a COVID-19 vaccine by 15th March 2020 | NIMS |
| Age | Third-order polynomial | 2011 Census |
| Sex | Female, Male | 2011 Census |
| Ethnicity | White British, Bangladeshi, Black African, Black Caribbean, Chinese, Indian, Mixed, Other, Pakistani, White other | 2011 Census |
| Religious affiliation | Christian, Buddhist, Hindu, Jewish, Muslim, No religion, Other Religion, Religion Not Stated, Sikh | 2011 Census |
| Region | Dummy variables representing region of residence | 2019 NHS PR |
| Rural urban classification | Urban, rural | 2019 NHS PR |
| Index of Multiple Deprivation (IMD) | Dummy variables representing quintiles of deprivation | 2019 NHS PR |
| Household tenure | Own, social rented, private rented, other | 2011 Census |
| Level of highest qualification | Degree, A-level or equivalent, GCSE or equivalent, no qualification, other | 2011 Census |
| Disability | Non-disabled , disabled (limited a little), disabled (limited a lot) | 2011 Census |
| Body Mass Index (kg/m<sup>2</sup>) | < 18.5, 18.5 – 25, 25 to 30, >= 30, missing | GPES |
| Chronic kidney disease (CKD) | No CKD, CKD3, CKD4, CKD5 | GPES |
| Learning disability | No learning disability, Down's Syndrome, other learning disability | GPES |
| Cancer and immunosuppression | Dummies for blood cancer, solid organ transplant , Prescribed immunosuppressant medication by GP , Prescribed leukotriene or long-acting beta blockers, Prescribed regular prednisolone , | GPES |
| Other conditions | Diabetes, Chronic obstructive pulmonary disease (COPD), Asthma , Rare pulmonary diseases , Pulmonary hypertension or pulmonary fibrosis , Coronary heart disease , Stroke , Atrial Fibrillation , Congestive cardiac failure , Venous thromboembolism , Peripheral vascular disease , Congenital heart disease , Dementia , Parkinson's disease , Epilepsy , Rare neurological conditions , Cerebral palsy , Severe mental illness (bipolar disorder, schizophrenia, severe depression), Osteoporotic fracture , Rheumatoid arthritis or Systemic lupus erythematosus , Cirrhosis of the liver | GPES/HES |

